# Magnetoencephalography reveals altered synaptic gain linked to functional impairment in adolescents at familial risk for schizophrenia and bipolar disorder

**DOI:** 10.64898/2026.11.20.26368386

**Authors:** Oskar Hougaard Jefsen, Maja Gregersen, Sinnika Birkehøj Rohd, Ron Nudel, Daniel J. Hauke, Anette Faurskov Bundgaard, Andreas Færgemand Laursen, Lotte Veddum, Merete Birk, Kathrine Skak Madsen, William Baaré, Kit Melissa Larsen, Hartwig Roman Siebner, Torben Lund, Leif Østergaard, Nicoline Hemager, Anne Amalie Elgaard Thorup, Merete Nordentoft, Aja Neergaard Greve, Ole Mors, Karl Friston, Yury Shtyrov, Martin Dietz

## Abstract

**BACKGROUND:** Auditory oddball responses, including the mismatch negativity (MMN), are impaired in patients with schizophrenia (SZ) and bipolar disorder (BP). Biophysical modelling of cortical microcircuit function and effective connectivity implicates reduced synaptic gain of excitatory pyramidal cells in patients with SZ, but whether similar alterations are present in adolescents with familial or genetic risk remains unknown.

**METHODS:** We studied 15-year-old adolescents enrolled in The Danish High Risk and Resilience Study – VIA 15. We measured auditory oddball responses in 59 adolescents at familial high risk for SZ (FHR-SZ), 38 adolescents at familial high risk for BP (FHR-BP), and 75 population-based controls (PBC) using magnetoencephalography (MEG) during a roving oddball paradigm. We investigated sensor- and source-level evoked responses and modelled effective connectivity using dynamic causal modelling (DCM). We examined the impact of familial and polygenic risk for SZ and BP, and associations with global level of functioning and auditory psychotic experiences.

**RESULTS:** Sensor- and source-level analyses did not detect statistically significant differences in oddball responses between adolescents at FHR-SZ and FHR-BP compared with PBC. However, DCM analyses revealed reduced oddball-dependent gain of pyramidal cells in an empirically determined left frontal source in FHR-SZ and FHR-BP adolescents compared with PBC, and this was associated with lower levels of functioning across groups.

**CONCLUSIONS:** Biophysical modelling of auditory oddball responses identifies surprise-related differences in effective connectivity in adolescents at FHR-SZ and FHR-BP. These findings suggest that microcircuit dysregulation, particularly in frontal synaptic gain, may serve as an early marker of vulnerability.

## Introduction

Schizophrenia (SZ) and bipolar disorder (BP) are severe psychiatric disorders with overlapping clinical and genetic features (1–3) typically diagnosed in early adulthood (4). Premorbid neural biomarkers may provide insights into underlying mechanisms and offer opportunities for earlier detection and intervention (5, 6). One candidate biomarker is the auditory oddball response measured with electroencephalography (EEG) or magnetoencephalography (MEG)(7), which offer exquisite millisecond-scale temporal resolution to characterise neural processing (8). In oddball paradigms, repeated ‘standard’ stimuli are interrupted by an unexpected deviant stimulus. The difference between deviant and standard responses constitutes the oddball response, which includes the mismatch negativity (MMN), an evoked response component peaking around 100-250 milliseconds. The MMN is reliably reduced in patients with SZ and BP and has been shown to be predictive of future conversion to psychosis in clinical high-risk groups (5, 9, 10). Twin studies further demonstrate high heritability of MMN reductions, overlapping with the heritability of SZ and BP (11–14). Furthermore, adult first-degree relatives of patients with SZ exhibit reduced MMN (9), suggesting that MMN reductions may represent an intermediate phenotype leading from genetic risk to illness. However, studies on children and adolescents — born to parents with SZ or BP — are sparse, limiting our understanding of whether, and how, MMN reductions are expressed in individuals at familial risk (15–17). Characterising these alterations in youth at familial risk may provide novel mechanistic insights into the shared and distinct developmental pathophysiology of schizophrenia and bipolar disorder.

The MMN is linked to several core mechanisms of psychosis, including aberrant predictive coding within auditory networks (18, 19), grey-matter loss (20), *N*-methyl-*D*-aspartate (NMDA) receptor hypofunction (21), and synaptic deficits in excitatory pyramidal cells (22, 23). Advances in biophysical modelling of cortical microcircuit dynamics offer new tools to interrogate these mechanisms non-invasively. Dynamic causal modelling (DCM) of EEG and MEG data uses generative modelling to fit models of mass neuronal network activity to empirical data, enabling formal comparison of competing hypotheses and estimation of effective connectivity within and between cortical sources. Previous DCM studies of MMN deficits have reported altered extrinsic (between-region) and intrinsic (within-region) connectivity in patients with SZ (22–24). Using DCM, Adams et al. (2022) found consistent evidence, across EEG and functional magnetic resonance imaging (fMRI) paradigms, for reduced synaptic gain of superficial pyramidal cells in bilateral inferior frontal gyri (IFG) among patients with SZ, a finding that may reflect the loss of synaptic density observed in post-mortem and positron emission tomography (PET) studies (25–28) of patients with SZ. Moreover, the gain of superficial pyramidal cells plays an important role of predictive coding theories of brain functioning, determining the precision (weighting) of prediction errors in cortical hierarchies, such that lower gain reduces the influence of sensory evidence on higher-level inference (29, 30). Altered synaptic gain therefore provides a mechanistic link between neurophysiology, cortical synaptic changes, and predictive coding accounts of psychosis and underscores the importance of studying these circuits developmentally (31). Similar methods have, to our knowledge, so far not been applied to data from patients with BP. And importantly, no study has mapped whether these circuit-level mechanisms are evident during adolescence, underscoring a crucial need for investigations in youth at familial high risk of SZ and BP.

*The Danish High-Risk and Resilience Study – the VIA cohort* (32) provides a unique opportunity to investigate auditory brain connectivity in offspring at familial high risk (FHR) for SZ (FHR-SZ) and BP (FHR-BP) along with population-based controls (PBC). Previous investigations included EEG recordings from the Copenhagen site (n = 173), which revealed alterations in effective connectivity in 11-year-old children at FHR-SZ and FHR-BP relative to PBCs (17). Here, we analyse MEG data from the Aarhus site of the most recently completed wave of the VIA study, leveraging the high temporal and spatial resolution offered by MEG. Our primary aim is to test whether familial and genetic liability to SZ and BP are associated with altered auditory effective connectivity and synaptic gain in adolescence. As a secondary aim, we examine associations with auditory psychotic experiences (33) and global level of functioning.

## Methods

### Participants

*The Danish High Risk and Resilience Study – the VIA cohort* is a nationwide longitudinal cohort study of 522 children at FHR-SZ (N = 202), FHR-BP (N = 120), and PBCs (N = 200). Nationwide Danish registers (34, 35) were used to identify parents with SZ or BD, and parents with neither of those disorders (PBCs). We recruited children from these families, matching PBC children to FHR-SZ children on age, sex, and municipality. We assessed the children every fourth year from the age of 7 (32, 36, 37). Assessments took place at one of two sites, in Aarhus and Copenhagen. We acquired MEG data at the Aarhus-site in the third wave (VIA 15) from mid-2021 to mid-2024. This study represents data from age 15, the VIA 15 study, which was approved by the National Committee on Health Research Ethics (H-20067908) and the Danish Data Protection Agency (P-2019-273). The genetic part of the VIA project was added as part of the *Arv og Miljø* protocol (H-B-2009-026). Supplementary Figure S1 displays the selection and retention thorough assessment waves in a flowchart.

### Assessments

We performed detailed assessments of all participants using a broad test battery as described in published protocols (32, 36, 37). Here we focused on the following measures. We rated the participants’ level of functioning in the previous month using the semi-structured interview Children’s Global Assessment Scale (CGAS)(from 1-100; higher scores representing better functioning)(38). We assessed parental level of functioning using the Personal and Social Performance (PSP) scale (from 1-100; higher scores representing better functioning)(39). Parents reported their offspring’s behavioural-emotional problems using the parent-reported Child Behaviour Checklist for ages 6-18 (CBCL)(40)(from 0-236; higher scores representing more problems). We examined the 4-year prevalence of auditory psychotic experiences (PE) using the Kiddie Schedule for Affective Disorders and Schizophrenia – Present and Lifetime version (K-SADS-PL) semi-structured diagnostic interview (41)(see Supplement for details).

### Polygenic risk scores

All participants were of European ancestry. The quality control and imputation procedure have been described previously (42). For this study we generated polygenic risk scores (PRS) for SZ and BP using updated summary statistics from large genome-wide association studies (GWAS) on Europeans, excluding Danish samples to avoid sample overlap (43, 44). PRSs were generated using PRSice v2.3.5 (45) using a *p*-value threshold of pT = 1 (see Supplement for details).

### Auditory oddball paradigm

Participants were seated in a magnetically shielded and sound-attenuated room and received auditory stimuli binaurally through silicone tubes connected to in-ear earpieces. The stimuli were controlled by Presentation software (Neurobehavioral Systems, Inc., Albany, CA, USA). Prior to data acquisition, we determined participants’ hearing threshold using a manual staircase algorithm. During data acquisition, participants watched a silent film and were instructed not to attend to the auditory stimuli. We presented sequences of pure tones (45 dB above hearing threshold; 75 ms in duration, including 5 ms fade-in and 5 ms fade-out) between 500 and 1000 Hz at 100 Hz intervals, which changed in a roving oddball pattern (18)(Figure 1A) with a stimulus onset asynchrony of 500 ms. The number of repetitions at the same frequency varied randomly between 2 and 10. A total of 200 sequences were presented. The roving paradigm ensures independence between the oddball contrast (deviants versus standards) and the stimuli’s physical properties (frequencies). During data acquisition, participants watched a silent film and were instructed not to attend to the auditory stimuli.

### Neuroimaging

We collected MEG data at the Center of Functionally Integrative Neuroscience (CFIN, Aarhus University / Aarhus University Hospital) using a Neuromag TRIUX 306-channel MEG system (MEGIN Inc., Helsinki) at a sampling rate of 1000 Hz. We used continuous head position indicators (cHPI) to monitor head position. We recorded cardiac activity (electrocardiogram, ECG), and vertical and horizontal eye movements (electrooculogram, EOG) with surface electrodes. Approximately one hour after MEG acquisition, we collected anatomical T1-weighted images (see Supplement) for participant-specific source localisation of MEG activity (i.e., construction of an electromagnetic forward model for each participant).

### MEG preprocessing

Preprocessing steps are described in detail in the Supplement. In short, MEG data were preprocessed using the temporal extension of the spatiotemporal signal space separation technique (tSSS)(46) and subsequently analysed using Statistical Parametric Mapping toolkit (SPM12, r7771). Data were filtered (0.5-30 Hz), downsampled to 250 Hz, and divided into 500-ms epochs (-100 to 400 ms relative to stimulus onset). We rejected artefacts using a simple threshold algorithm (see Supplementary). We performed robust averaging over epochs, defining the first tone in each sequence as ‘deviants’ and the 5^th^-10^th^ tones as ‘standards’. We subtracted the averaged standard response from the deviant response, yielding the ‘oddball response’ per subject, and converted the magnetometer data (0 to 400 ms) to three-dimensional (scalp x time) images for statistical analysis.

### Source localization

The acquisition and processing of structural MRIs is described in the Supplement. We constructed forward models with cortical meshes generated from structural MRIs (individual or template), co-registered with MEG recordings using fiducial markers (nose and preauricular points) and head shape-points, based on a single-shell head model. We inverted the forward models using multiple sparse priors (MSP)—under group constraints—to yield participant-specific source estimates and converted the evoked source-level activity into 3D images for statistical analysis. Across all participants, we identified sources showing oddball effects, i.e., the evoked response during deviants compared to standards (paired *t*-test, FWE < 0.05), and obtained coordinates for source priors in subsequent DCM. Finally, we extracted virtual electrode data from these sources (Figure 1C) and converted these to one-dimensional images for statistical analysis.

### Dynamic causal modelling

Dynamic causal modelling (DCM) is a Bayesian framework for inferring latent neural mechanisms underlying observed time-series (SPM12, r7771). We specified a hierarchical auditory network based on previous fMRI, EEG, and DCM studies of auditory oddball responses (18, 22–24, 47–50) as well as the present source localisation of oddball effects. This network included six sources: bilateral primary auditory cortices, posterior superior temporal gyri (STG), and precentral gyri (PCG), connected via feedforward, feedback, and intrinsic self-inhibitory connections (Figure 2A). Each source was modelled using a modified canonical microcircuit (22, 51) with four neural populations—superficial pyramidal (sp), inhibitory interneurons (ii), spiny stellate (ss), and deep pyramidal cells (dp) —featuring recurrent sp–ii connections and an excitatory sp-dp connection, consistent with cortical anatomy (Figure 2D), and in line with previous DCM studies (22, 29, 51). Among the intrinsic connections, only the self-inhibitory sp-sp connection (inverse synaptic gain) was allowed to vary between conditions, given its central role in predictive coding theory (29, 30). Full models (estimating all feedforward, feedback, and intrinsic connections) were inverted using Variational Laplace (52) and refined—at the group level—via iterative parametric empirical Bayes (PEB) until convergence (53)(see Supplementary Methods and Supplementary Figure S5). This application of PEB uses group-level posteriors as individual-level priors, rescuing individual DCMs from local minima, without biasing subsequent group comparisons.

### Sensitivity analysis

To illustrate the mechanistic contributions of DCM condition-dependent (*B*) parameters to sensor-level data, we fitted a DCM to the PBC grand-mean response and simulated how increases/decreases to ensuing parameters affected sensor-level responses. We plotted the simulated sensor-level responses at sensors showing scalp-level group differences or associations with continuous measures (e.g. CGAS)(p < 0.001, uncorrected).

### Statistical analysis

Statistical analyses were performed in MATLAB R2024b (54). We compared participant characteristics between groups using Pearson χ^2^, analysis of variance (ANOVA) or Kruskal-Wallis’s test, and subsequent pair-wise comparisons employing t-tests or Wilcoxon rank sum tests. As dropout analyses, we compared VIA 15 MEG study participants versus non-participants on their level of functioning and of emotional and behavioural problems at the first and current study wave (VIA 7 and 15) using t-tests or Wilcoxon rank sum tests. We investigated scalp- and source-level oddball responses in PBCs using statistical parametric maps (SPMs) with one-sample *t*-tests, and compared scalp- and source-level oddball responses between groups using analysis of covariance, adjusting for sex and age. All sensor- and source-level analyses were thresholded using random field theory to control the family-wise error (FWE) rate at *p* < 0.05. We plotted the grand-mean oddball response for each group from the sensor nearest the maximum of the oddball effect, for visualisation. We used parametric empirical Bayes (PEB) (55) to perform group analyses of DCMs (56). We first tested for oddball effects on effective connectivity in the PBCs alone. We then tested for group differences in condition-dependent connectivity, in two separate models, one comparing FHR-SZ vs. PBC, and one comparing FHR-BP vs. PBC, both adjusting for sex and age. We then tested for associations between PRS for SZ or BP and condition-dependent connectivity, adjusting for sex, age, and FHR status. Finally, we tested for associations with auditory psychotic experiences (+/-) and level of functioning (CGAS), adjusting for sex, age, and FHR status. We used Bayesian model reduction (BMR) to search over all nested models within the full models (i.e., a post-hoc search), followed by Bayesian model averaging (BMA), which evaluates group-level posteriors from all models, weighted by their respective model-evidence (55). We plotted means and credible intervals for all parameters with a posterior probability > 0.95 (55).

## Results

### Included participants and clinical characteristics

We acquired data from 174 adolescents, of whom 172 provided usable MEG data (one participant was excluded due to excessive movement in MEG, and another due to faulty trigger data). Supplementary Figures S1 and S2 display a flowchart of participant recruitment and magnetoencephalography data quality measures, respectively. Table 1 describes the clinical characteristics of the final sample. Adolescents at FHR-SZ and FHR-BP had lower levels of global functioning, lower parental levels of social functioning, and higher levels of behavioural and emotional problems than PBC subjects (Table 1). The analysed MEG sample represented 33% of the original VIA 7 sample (N=522) with equal proportions from each group (Pearson χ² = 2.6, *p* = 0.27). The VIA 15 MEG study participants and non-participants showed similar levels of functioning and emotional and behavioural problems both at age 7 and 15 (Tables S1 and S2). Individual DCMs were successfully fitted for all participants; but two participants failed inversion during iterative fitting (during the second iteration) and were excluded from group-level analysis of DCM parameters.

### Oddball processing

PBCs exhibited significant condition effects (DEV vs. STD) on scalp-level event-related fields (ERFs) (Figure 1B, Table S3-4), bilaterally, at both early (peaking at 152 ms) and late (368 ms) time points. We localised auditory oddball effects (DEV vs. STD), over all subjects, to multiple cortical sources (Figure 1C-D, Table S5), including bilateral A1, STG, and frontal sources covering the precentral gyri (PCG), pars opercularis of the IFG, and opercular cortices (from here on referred to as ‘PCG’). Bayesian model comparison favoured a cortical network based on empirical sources, rather than on Garrido et al. (19) (Figure 1E), and empirical sources were thus used for DCM. In PBC, oddball processing (DEV vs. STD) was associated with decreased self-inhibition (increased gain) in R-A1, increased feedforward connectivity from bilateral A1 to STG, decreased feedback from R-STG to R-A1, and decreased L-PCG self-inhibition (increased gain) (Figure 1F, Table S6).

**Figure 1.**
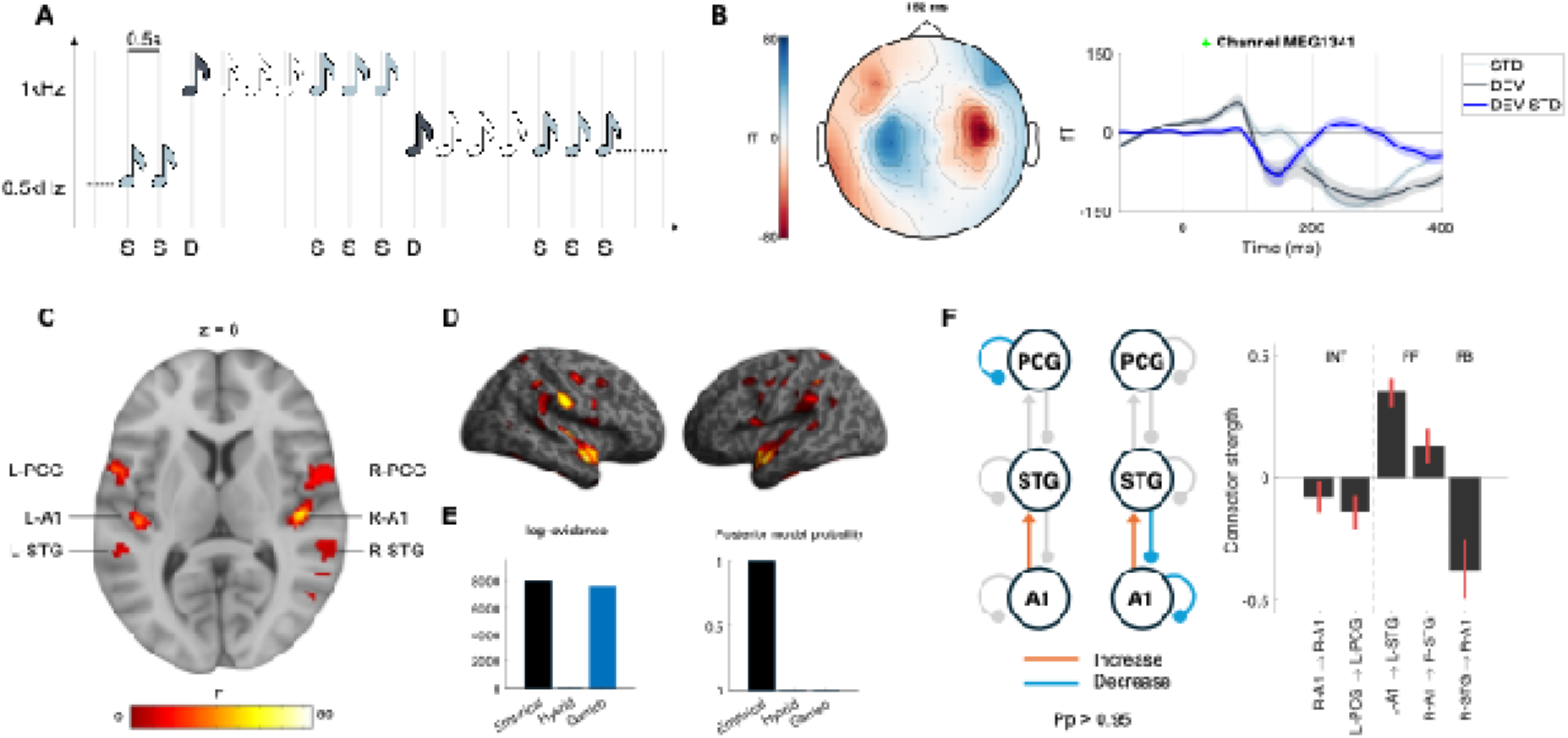
Auditory oddball responses. A) Roving auditory oddball paradigm containing standards (S) and deviants (D). B) Scalp topography at time = 152 ms (peak oddball effect) in population-based controls (PBC) and sensor-level event-related fields for standards (STD), deviants (DEV), and their contrast (DEV-STD) at a single channel near the peak oddball effect in population-based controls (PBC), C) Display of sources differentially activated by deviants vs. standards (p < 0.05), at a single section (see more sections in Supplementary Figure S4). D) Cortical surface renderings of source localisation contrast (DEV vs. STD). E) Bayesian model comparison of dynamic causal models using different source coordinates: either fully Empirical, derived from Garrido et al. 2007, or a hybrid with empirical primary auditory cortex (A1) and superior temporal gyrus (STG) sources plus the inferior frontal gyrus (IFG) sources based on Garrido et al. 2007. F) Oddball-dependent effective connectivity (DEV vs. STD) in PBCs only, with modulation of both feedforward (FF), feedback (FB), and intrinsic (INT) effective connectivity. Only effects with a posterior probability (Pp) > 0.95 are shown. Error bars reflect 95% Bayesian credible intervals.

### Associations between familial risk and oddball processing

We detected no statistically significant differences between FHR-groups and PBCs in scalp- or source-level ERFs at *p* < 0.05, FWE-corrected for multiple comparisons (see Supplementary Figure S3 for visualization). However, using an uncorrected threshold of *p* < 0.001, we observed differences between FHR-SZ and PBC (Supplementary Table S7 and Figure S3), but not between FHR-BP and PBC. In contrast, DCM analyses revealed strong evidence (i.e., posterior probability (Pp) > 0.95) for alterations in oddball-dependent intra-cortical connectivity between groups. Adolescents at FHR-SZ showed increased L-PCG self-inhibition for DEV vs. STD, indicating lower synaptic gain during oddball processing (Figure 2, Table S8). Similarly, adolescent at FHR-BP showed increased L-PCG self-inhibition (lower synaptic gain), decreased L-STG self-inhibition (higher synaptic gain), and increased R-A1 self-inhibition (lower synaptic gain) (Figure 2, Table S9). In a combined analysis of both high-risk groups compared to PBCs, adolescents at FHR-SZ or FHR-BP showed increased L-PCG self-inhibition (lower synaptic gain) and decreased L-STG self-inhibition (higher synaptic gain) (Table S10, Supplementary Figure S6).

**Figure 2.**
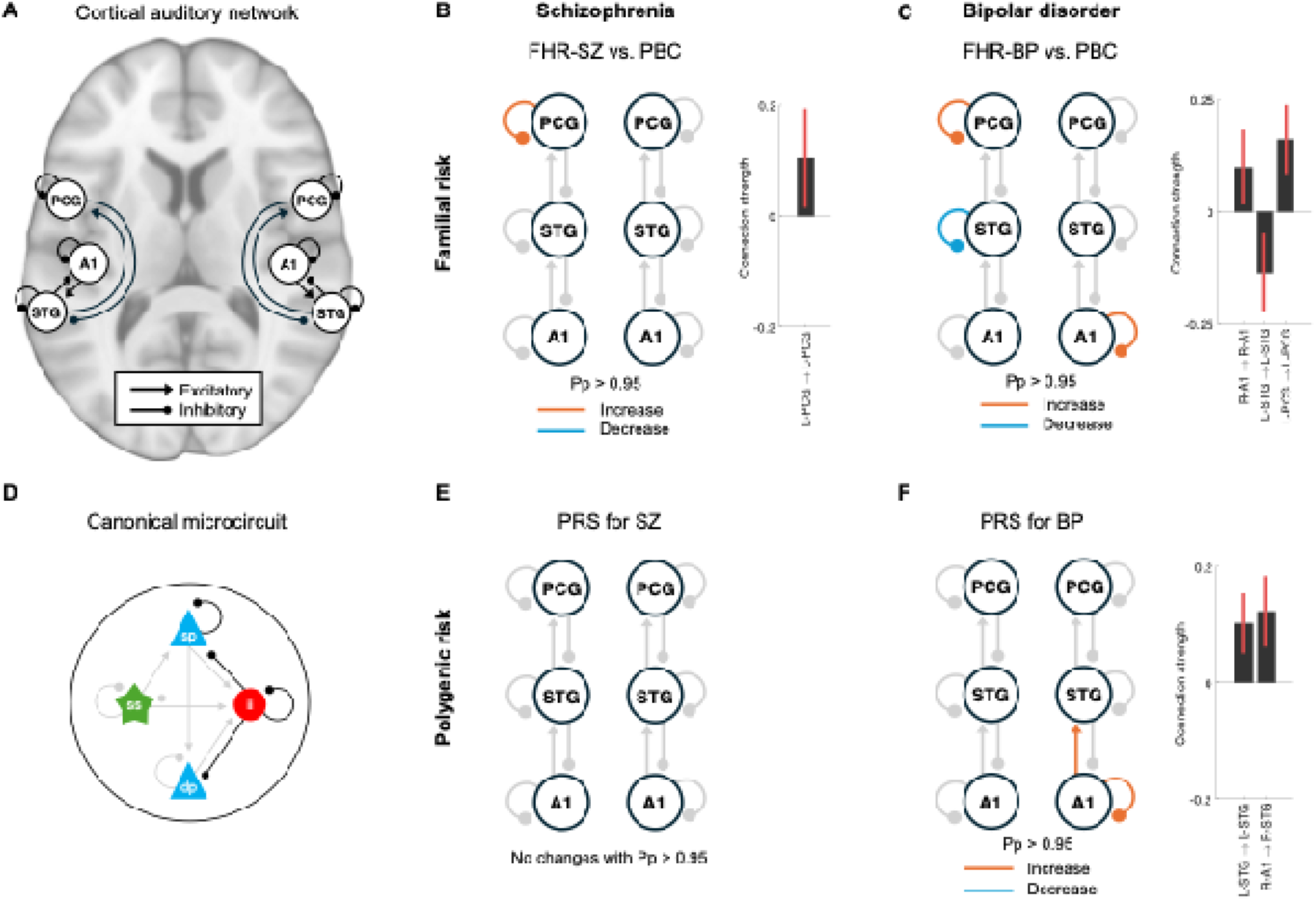
Associations between familial and polygenic risk of schizophrenia and bipolar disorder and oddball-dependent connectivity. A) Auditory network used in dynamic causal modelling (DCM), including bilateral primary auditory cortex (A1), superior temporal gyrus (STG), and precentral gyrus (PCG). Sources are connected via feedforward (excitatory) and feedback (inhibitory) connections. The gain of each source is parameterised as the self-inhibitory connection on superficial pyramidal cells. B) Oddball-dependent effective connectivity in adolescents at familial high risk of schizophrenia (FHR-SZ) versus population-based controls (PBC). C) Oddball-dependent effective connectivity in adolescents at familial high risk of bipolar disorder (FHR-BD) versus population-based controls (PBC). D) Canonical microcircuit (CMC) used in DCM. The circuit is comprised of superficial pyramidal (sp) cells, spiny stellate (ss) cells, inhibitory interneurons (ii), and deep pyramidal (dp) cells. Intrinsic connections marked in black are optimised/estimated during model inversion, while those marked in grey are fixed to their prior. E) Effect of polygenic risk of schizophrenia (SZ) on oddball-dependent effective connectivity across the entire sample. F) Effect of polygenic risk of bipolar disorder (BD) on oddball-dependent effective connectivity, across the entire sample. Only effects with a posterior probability (Pp) > 0.95 are shown. DEV = deviants. STD = standards. Error bars represent 95% Bayesian credible intervals.

### Associations between polygenic risk and oddball processing

PRSs for SZ or BP were not associated with oddball ERFs at the sensor or source level. Polygenic risk for SZ was not associated with oddball-dependent connectivity (Figure 2, Table S11), while higher polygenic risk for BP was associated with increased R-A1 self-inhibition (lower synaptic gain) and stronger feedforward connectivity from R-A1 to R-STG (Figure 2, Table S12).

### Associations between auditory psychotic experiences and oddball processing

In the MEG sample, nine participants reported at least one auditory psychotic experience within the past 4 years (PE+), whereas 163 did not (161 in PEB analyses)(PE-). No sensor- or source-level ERF differences were observed between PE+ vs. PE-participants. However, DCM analysis showed strong evidence (Pp > 0.95) for an association of auditory psychotic experiences and a decrease of L-STG self-inhibition (higher synaptic gain) (Figure 3A, Table S13).

**Figure 3.**
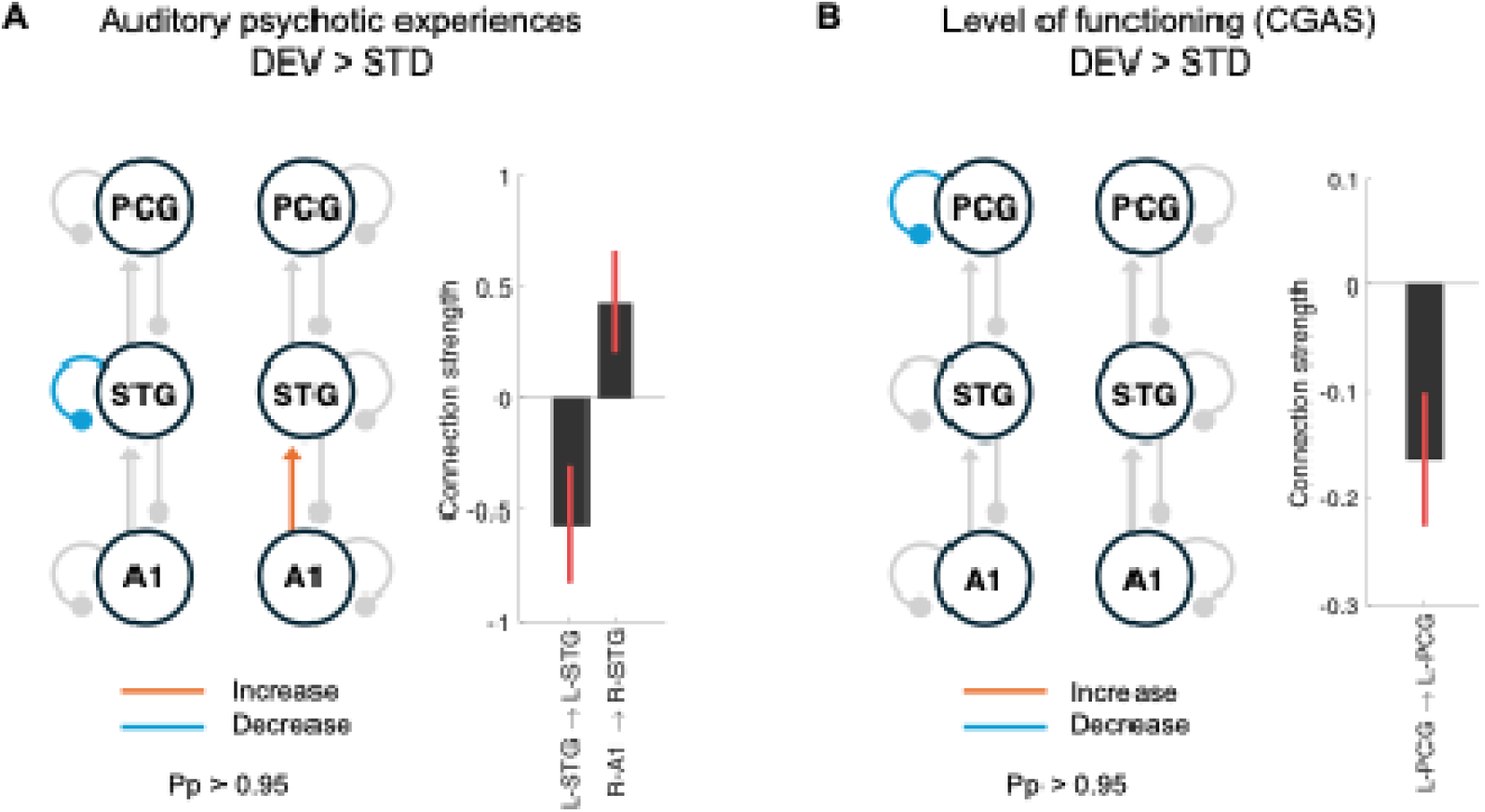
A) Oddball-dependent connectivity in adolescents with (n = 9) vs. without (n = 161) auditory psychotic experiences. B) Association between level of functioning (Children’s Global Assessment Scale (CGAS); higher score = better functioning) in adolescents across the total sample (n = 170). Only effects with a posterior probability (Pp) > 0.95 are shown. Error bars represent 95% Bayesian credible intervals. DEV = deviants. STD = standards. PCG = precentral gyrus. STG = superior temporal gyrus. A1 = primary auditory cortex.

### Associations between level of functioning and oddball processing

We detected no statistically significant associations between sensor- or source-level ERFs and CGAS at *p* < 0.05, FWE corrected for multiple comparisons. However, using an uncorrected threshold of *p* < 0.001, we observed a positive association between CGAS and sensor-level ERF strength at 184 ms over the right frontal region and a negative association between CGAS and sensor-level ERF strength at 96 ms over the left frontal region (Supplementary Table S14). DCM analyses indicated strong evidence that higher CGAS (better functioning) was associated with lower self-inhibition (higher synaptic gain) within L-PCG (Figure 3B, Table S15). This association remained after controlling for high-risk status.

### Simulations reproduce sensor-level effects of left precentral gyrus self-inhibition

Following the observed differences in L-PCG self-inhibition in FHR-SZ and FHR-BP compared with PBC, and associations with level of functioning, we tested whether simulated changes in this parameter would reproduce the sensor-level differences observed empirically. This revealed that an increase in L-PCG self-inhibition (the alterations in FHR-SZ and FHR-BP) reproduced the more negative peak in FHR-SZ compared to PBC at 224 ms (Figure 4). Moreover, a decrease in L-PCG self-inhibition (the alteration in those with a higher level of functioning) reproduced the amplitude-reduction observed at 136 ms with a higher level of functioning (Figure 4).

**Figure 4.**
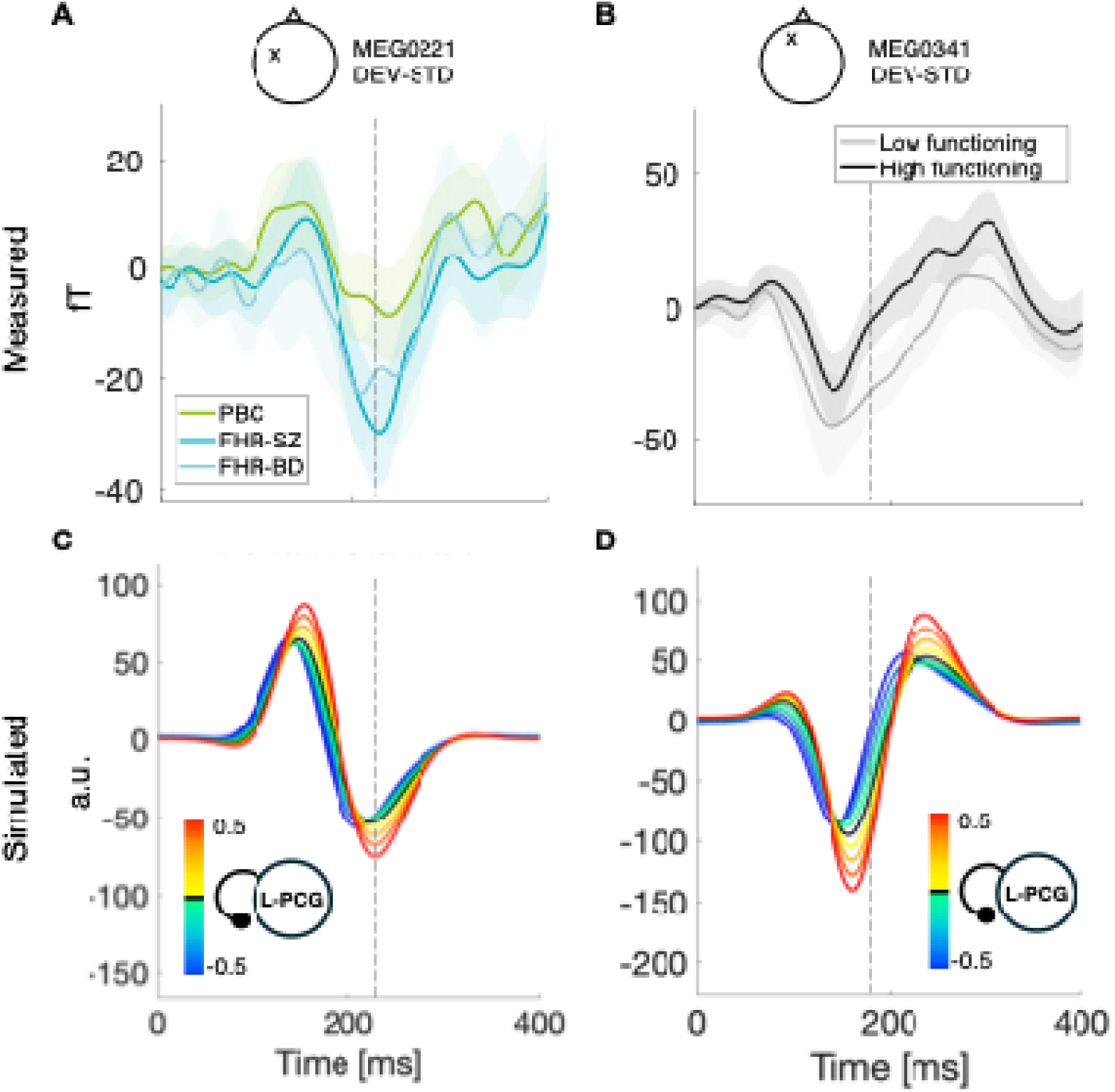
Sensitivity analyses show the measured (top [A and B]) and simulated (bottom [C and D])) oddball-responses (deviants minus standards, DEV-STD) at channels MEG0221 (left [A and C]) and MEG0341 (right [B and D]). These channels were selected as those nearest trend-level associations observed at the scalp levels for the FHR-SZ vs. PBC contrast and for the level of functioning. Dashed lines indicate the time points for the trend-level associations (A/C: 224 ms; B/D: 184 ms). Left precentral gyrus (L-PCG) self-inhibition was increased (yellow-red lines) or decreased (green-blue lines) in simulations, yielding altered sensor-level responses. fT = femtotesla. a.u. = arbitrary units.

## Discussion

Using MEG and DCM, we investigated cortical auditory evoked responses in adolescents at FHR-SZ or FHR-BP, compared with PBCs, and modelled the effects of familial and polygenic risk. Sensor- and source-level analyses indicated no significant associations between familial or polygenic risk and classical oddball evoked responses (MMN) at the age of 15. Using the DCM framework, biophysical modelling revealed evidence for inferior frontal cortical alterations in auditory effective connectivity in adolescents at FHR-SZ and FHR-BD compared with PBC, associations with PRS for SZ, level of global functioning, and suggestive links to auditory psychotic experiences.

Biophysical modelling of the MEG data revealed group differences in effective connectivity. These differences included a reduced gain of excitatory pyramidal cells in the L-PCG in adolescents at FHR-SZ and FHR-BP. A similar effect was found in adolescents with a lower level of functioning, also present after adjustment for familial high-risk status. These effects are consistent with the findings by Adams et al. (22) and Ranlund et al. (23), who reported lower oddball-dependent gain of pyramidal cells in IFG in patients with SZ using DCM analyses of EEG data. Yet, other DCM-EEG studies found no association with IFG synaptic gain (17, 24). Larsen et al. previously modelled EEG data from another subsample of the VIA cohort at age 11 (the VIA 11 study, Copenhagen site), assessed with a classical double-deviant auditory oddball paradigm (17). They found reduced feedforward connectivity from L-STG to L-IFG in children at familial high risk of SZ or BP and a modulation of L-IFG self-inhibition, when compared with PBCs. Importantly, direct comparison between studies is complicated by the differences in age and the use of different neuronal models with different parameterisation of some biophysical model features (57).

An important difference from the previous DCM studies is in the exact location of frontal sources modelled. Most previous DCM studies did not perform independent source localization but used source coordinates based on the stereotactic coordinates reported by Rademacher et al. (58) and fMRI coordinates reported by Opitz et al. (49), which included the L/R STG and R-IFG (19, 22, 23, 59). Similar to recent large MEG studies (60, 61), we did not detect sources in the IFG, which may be due to the lower sensitivity of MEG to radial sources (relative to EEG; but see (62). However, we detected significant oddball effects in adjacent bilateral areas covering ventral PCG, the frontal operculum, and a small part of the IFG pars opercularis; roughly 18 mm posterior to the IFG coordinates typically used. The PCG has, like the IFG, previously been implicated in higher-order auditory processing (63–65) and Bayesian model comparison favoured a cortical network with these frontal sources rather than the IFG coordinates used in earlier studies, supporting the modelling of these regions in the present study.

The absence of significant associations between familial risk of schizophrenia and classical sensor- or source-level ERFs is in line with previous studies (15–17), and raises the question of when the clear ERF/ERP alterations seen in adult first-degree relatives (9) emerge. As previously shown (66–68), the maturation of the auditory mismatch negativity continues well into adolescence or even adulthood. Trait-dependent deficits in first-degree relatives may, speculatively, depend on this maturation.

Our analyses also revealed circuit signatures of subclinical psychotic phenomena, including reduced self-inhibition of the L-STG (higher synaptic gain) in individuals with auditory psychotic experiences. These preliminary results need to be interpreted with caution, given the small sample size with only nine persons reporting auditory psychotic experiences. However, these results align with previous DCM studies linking positive psychotic symptoms in patients with higher cortical excitability (increased gain)(59). Critically, this disinhibition of the L-STG, which includes Wernicke’s area for receptive language (69), is biologically consistent with the elevated blood-oxygenation-level-dependent responses and glutamate levels previously observed in this area in hallucinating patients with schizophrenia (70, 71).

### Limitations

Although this study leverages a large well-characterized sample, high-precision neurophysiological recordings, polygenic analysis, and advanced statistical modelling, several limitations warrant consideration. First, we were unable to study the effects of manifest psychosis, as too few (<5) study participants had received such a diagnosis at age 15 (VIA 15)(72). Moreover, our study was cross-sectional, hindering longitudinal comparison between ‘converters’ to psychosis and ‘non-converters’. However, the ongoing VIA19 study (i.e., follow-up at age 19) will address this as more individuals are expected to have developed a psychotic disorder by this age. Second, the lack of significant group differences in sensor- and source-level analyses may appear counterintuitive given the strong evidence for group differences in the DCM results. This divergence is likely due to the specifics of the analyses, as the two methods test very different aspects: The former examines data points in time and space (sensor or source amplitudes over post-stimulus time), whereas DCM estimates parameters of a generative model of synaptic connections. Moreover, our simulations confirmed that DCM explained relevant data features, as changes to L-PCG self-inhibition successfully reproduced the timing and direction of the differences observed at the sensor level. Third, while we focused on the conventional deviant–standard contrast, future work could better characterise sensory learning during the roving paradigm by applying other methods (73, 74) that model all stimulus repetitions. Fourth, the observed alterations in synaptic gain may arise from multiple mechanisms, including changes in synaptic density (26), NMDA receptor hypofunction (75), and context-dependent inhibition (76). Our current analyses cannot distinguish among these, but future animal or human studies may help clarify their contributions. Fifth, our findings regarding auditory psychotic experiences were based on a small sample (n = 9 with PE) and are thus preliminary and require independent replication in more powered studies.

In summary, our findings demonstrate that combining MEG with biophysical modelling of neural population dynamics can non-invasively uncover circuit-level mechanisms underlying psychiatric vulnerability and disease. Our findings reveal how familial and genetic risk for schizophrenia and bipolar disorder manifest in adolescent auditory brain circuits, bridging genetic vulnerability with neural systems underlying psychosis.

## Data Availability

The data produced in the current study are not publicly available due to privacy and ethical restrictions.

## Acknowledgments

OHJ was funded by the Health Research Foundation of the Central Denmark Region (Grant no. R64-A3090-B1898) and the Lundbeck Foundation (Grant no. R484-2024-1581). KML was supported by the Lundbeck Foundation (Grant no. R322-2019-2311). The Danish High Risk and Resilience Study – VIA 15 was funded by The Lundbeck Foundation (Grant no. R277-2018-594 and R322-2019-2711), Novo Nordisk Foundation (Grant no. NNF20OC0060468), the Central Denmark Region, and the Mental Health Services of the Capital Region of Denmark. MD was supported by the Lundbeck Foundation (Grant no. R322-2019-2711). KF is supported by funding from the Wellcome Trust (Ref: 226793/Z/22/Z). YS was supported by the Lundbeck Foundation (Grant no. R427-2023-160).

We express our sincere gratitude to all participants and their families for their time and contribution to the study. We thank Johannes Reinau Windelborg Nielsen for assistance with MEG data acquisition, James Lubell for MEG technical support, Nanna Lawaetz Daugaard, Siri Væråmoen, and Henriette Norman Hansen for assistance with MRI data collection, and Nanna Weye for assistance with data management.

## CRediT authorship contribution statement

**Conceptualization:** OHJ, OM, KF, YS, MD. **Data Curation:** OHJ, MG, SBR, RN, TL, MD. **Formal analysis:** OHJ, MD. **Funding acquisition:** OHJ, NH, AAET, MN, ANG, OM, KF, HRS, YS, MD. **Investigation:** OHJ, AFB, AFL, MB, LV, MD. **Methodology:** OHJ, MG, SBR, KSM, WB, KML, HRS, KF, YS, MD. **Project administration:** OHJ, LV, MB, MD. **Resources:** KSM, WB, TL, LØ, NH, AAET, MN, ANG, OM. **Software:** KF, MD. **Supervision:** OM, KF, YS, MD. **Validation:** RN, MD. **Visualization:** OHJ, MD. **Writing – original draft:** OHJ. **Writing – review and editing:** All authors.

