## Supplementary Methods for "Magnetoencephalography reveals altered synaptic gain linked to functional impairment in adolescents at familial risk for schizophrenia and bipolar disorder"

to

**Short running title:** Modelling Oddball Responses in Familial Risk for Psychosis

**Keywords:** Schizophrenia, bipolar disorder, magnetoencephalography, dynamic causal modelling, familial high risk

**Corresponding author:** Oskar Hougaard Jefsen, MD, PhD. Psychosis Research Unit, Aarhus University Hospital. Palle Juul-Jensens Boulevard 175, 8200 Aarhus N, Denmark.

### Supplementary Methods

#### Assessment of auditory psychotic experiences

We performed the Kiddie Schedule for Affective Disorders and Schizophrenia – Present and Lifetime version (K-SADS-PL), a semi-structured diagnostic interview. We applied the K-SADS-PL psychosis supplement for all subjects, regardless of their responses to screening questions. A subsequent clinical conference with an experienced psychiatrist determined potential diagnosis/-es, the severity of each psychosis-like symptom (PLIKS) on a scale from 0-6. We categorized individuals with a severity rating of 2 or above on any auditory-domain item (1a, b, c, d, and f) as having auditory psychotic experiences (APE+), and all others as having no auditory psychotic experiences (APE-). Further details on these assessments are reported in Rohd et al. (1)

#### Polygenic risk scores

The generation of PRSs is described in detail elsewhere (2). DNA was obtained from saliva, genotyped on the Illumina PsychChip. to the genotype data underwent extensive quality control, and further markers were imputed prior to the computation of the PRSs. All remaining subjects after quality control were of similar genetic ancestry and clustered with European reference samples. We obtained summary statistics from the most recent Psychiatric Genomics Consortium (PGC) genome-wide association study (GWAS) meta-analyses restricted to individuals with European ancestry, with Danish individuals removed to avoid bias from sample overlap. The GWAS discovery sample for schizophrenia included 64,952 cases and 88,856 controls (3). The GWAS discovery sample for bipolar disorder included 55,912 cases and 740,590 controls (4). We generated PRSs using PRSice v2.3.5 using a p-value threshold of pT = 1.

#### Magnetic resonance imaging acquisition and processing

We obtained structural MRI on a Siemens Magnetom, 3.0 Tesla Skyra with a 32-channel head coil one hour after MEG recording. We acquired T1-weighted images of the whole head using a 3D magnetization-prepared rapid gradient echo (MP2RAGE) sequence (TR = 6500 ms, TE = 3.49 ms, flip angle = 0 degrees, field of view (FOV) = 260 x 260, 192 sagittal slices, voxel size 0.9 x 0.9 x 0.9 mm).

#### Magnetoencephalography preprocessing

We applied temporal signal space separation (tSSS) using Maxfilter (5), which helps remove external noise, detect bad channels, and uses continuous head position indicators (cHPI) to realign data after movement, and moving data to standard space across subjects. From cHPI data, we calculated the mean translational head movement, tracked every 200 ms (6).We performed all remaining processing and analysis using Statistical Parametric Mapping (SPM)(7). We performed high pass filtering (Butterworth, two-pass, fifth order) at 0.5 Hz, low-pass filtering (Butterworth, two-pass, fifth order) at 30 Hz, and downsampling of the data to 250 Hz. We epoched the data into 500 millisecond (ms) segments from -100 to 400 ms relative to stimulus onset (0 ms). We performed automatic artifact rejection using a simple threshold algorithm. Epochs with values above thresholds were rejected (for magnetometer data > 2500 femtoTesla (fT), or gradiometer data > 40 fT/millimeter) or marked as ‘bad’ (for standardized EOG data, z > 4, excision window 300 ms). Channels with more than 50% bad epochs were marked as bad. We then averaged data over epochs, using the first tone in each sequence as ‘deviants’, and the fifth through to the 10^th^ tone as ‘standards’, applying robust averaging which downweights potential outliers in the data. We subtracted the averaged standard response from the deviant response, yielding the ‘oddball’-response per subject, and converted the magnetometer-data, cropped from 0 to 400 ms in post-stimulus time, to three-dimensional (3D, scalp x time) images for statistical analysis.

#### Dynamic causal modeling

Dynamic causal modelling (DCM) is a type of Bayesian modelling scheme that allows inference on latent causes (i.e. the neural mechanisms) underlying observed data (here event-related fields, ERFs). We defined a hierarchical auditory network, based on our empirical MEG source localization (see Supplementary Figure S4 and Supplementary Tables) and previous fMRI, EEG and DCM-investigations on auditory oddball responses (8-15).

The network consisted of six sources with the following MNI coordinates (xyz):

- Left primary auditory cortex (L-A1):	[-41 -22 6]
- Right primary auditory cortex (R-A1):	[48 -16 6]
- Left posterior superior temporal gyrus (L-STG):	[-58 -42 10]
- Right posterior STG (R-STG):	[60 -42 10]
- Left precentral gyrus (L-PCG):	[-56 6 6]
- Right precentral gyrus (R-PCG):	[56 4 6]

Activity in each source was modelled using the canonical microcircuit (CMC), which consists of four neural populations: superficial pyramidal (sp) cells, inhibitory interneurons (ii), spiny stellate (ss) cells, and deep pyramidal cells (dp), as shown in Figure 2D. Modifying the original CMC, we added recurrent connections between sp and ii populations, and replaced the inhibitory sp → ss connection with an excitatory sp → dp connection. This modified CMC was similar to that used by Shaw et al. (16) and Adams et al. (14) and is closer to known cortical anatomy. We modelled the ‘baseline’ connectivity as the connectivity under standard stimuli (*A* parameters), as well as condition-specific connectivity as the oddball effect (deviants vs. standards)(*B* parameters). For each subject, we estimated four ‘baseline’ intrinsic connections (*G* parameters): sp → sp, sp → ss, ii → ss, ii → ii. The remaining were fixed to avoid overparameterization. Furthermore, the sp → sp self-inhibitory connection (inverse synaptic gain) was allowed to vary between conditions, due to its central role in predictive coding theory (17, 18). We applied the ‘IMG’ model and default inversion routines (Variational Laplace) (19). This method searches for the model with the highest log-model evidence, i.e., the highest negative variational free energy, *F*. We adopted an iterative, hierarchical empirical Bayesian fitting routine, in which individual DCMs were reinverted, using group-level posteriors as individual-level priors, until convergence – see Supplementary Methods for elaboration.

#### Iterative parametric empirical Bayesian inversion

Individual dynamic causal models (DCMs) are inverted using a gradient ascent on variational free energy, which approximates the log model evidence (20). A problem for model inversion, particularly when fitting complex models (as those used here), is the potential for multiple local maxima, which may hinder convergence and reliability. Individual DCMs may get stuck in locally optimal solutions, affecting both the posterior parameter estimates and the model evidence. This problem may be partially ameliorated by fitting full models (including all parameters) and subsequently applying Bayesian model reduction (BMR) to derive the model evidence for all reduced models (with some or all parameters removed) (21), although local minima may still lead the inversions astray. Another supplementary method that may reduce the problem is iterative parametric empirical Bayesian (PEB) inversion (22). In this method, individual DCMs are first inverted with standard priors. Then, a PEB analysis is performed on the individual DCMs to derive group-level posterior parameter estimates. After this step, each individual DCM is reinverted, using group-level posteriors as priors, and PEB is once again used to derive group-level posteriors. This scheme is iterated until the model converges. The log precision (inverse variance) is suggested as the primary criterion for convergence, which usually occurs after three or four iterations (22).

### Supplementary Figures

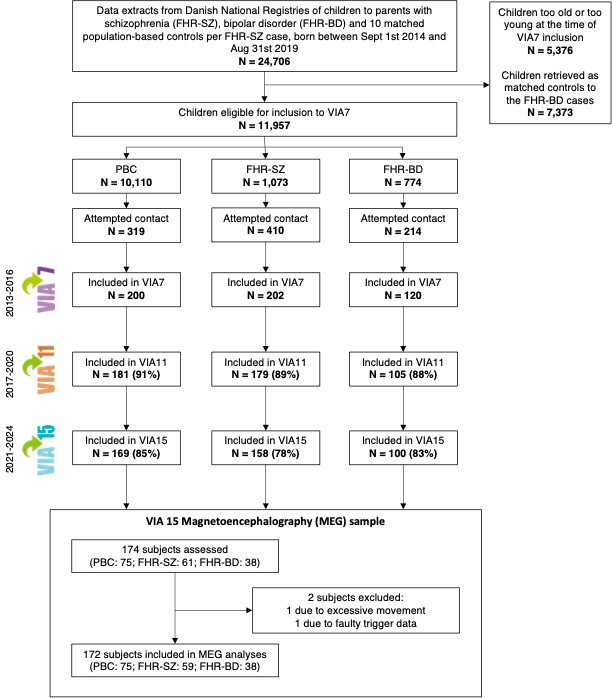

Supplementary Figure S1. Flowchart depicting recruitment to the study. Clinical and behavioural assessments were divided between two study sites: Aarhus and Copenhagen. The initial VIA7 protocol initially aimed to have all three groups matched on sex, age, and municipality, however this turned out to be unfeasible. Accordingly, some potential population-based control-children were excluded early in the process (top right corner). Percentages indicate proportion of the VIA7 sample. VIA15 magnetoencephalography only took place in Aarhus and primarily recruited individuals assessed in Aarhus, although some individuals assessed in Copenhagen participated in MEG in Aarhus. This cross-site MEG participation was primarily driven by practicalities, capacity, and participant preferences. Moreover, if the participant had been participated in magnetic resonance imaging (MRI) in Aarhus in VIA11, this participant was also booked for neuroimaging in Aarhus in VIA 15, to not change MRI-scanners between waves. FHR-SZ = familial high-risk of schizophrenia. FHR-BP = familial high-risk of bipolar disorder.

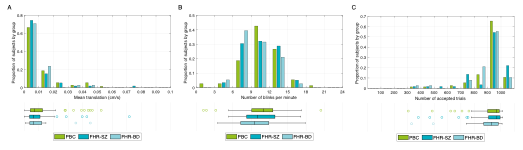

Supplementary Figure S2. Magnetoencephalography data quality measures. (A) Movement in scanner measured as mean translational movement (cm/s) calculated from Maxfilter log files. We analysed Maxfilter log files using scripts from <https://lsr-wiki-02.mrc-cbu.cam.ac.uk/meg/maxdiagnost> and calculated the mean of all time bins. Kruskal-Wallis’ test revealed no statistically significant differences between groups (p = 0.50). (B) The number of eye blinks per minute quantified using the artefact detection method described under Methods. Analysis of variance revealed no statistically significant differences between groups (p = 0.29). (C) The number of accepted trials after artefact rejection. Kruskal-Wallis’ test revealed no statistically significant differences between groups (p = 0.24). Top panels display histograms for the proportion of subjects within each study group, with histogram bars including the lower (not higher) edge values. Lower panels display box plots of the values by group with outliers (quartile one or three +/- 1.5 interquartile range) represented as single dots.

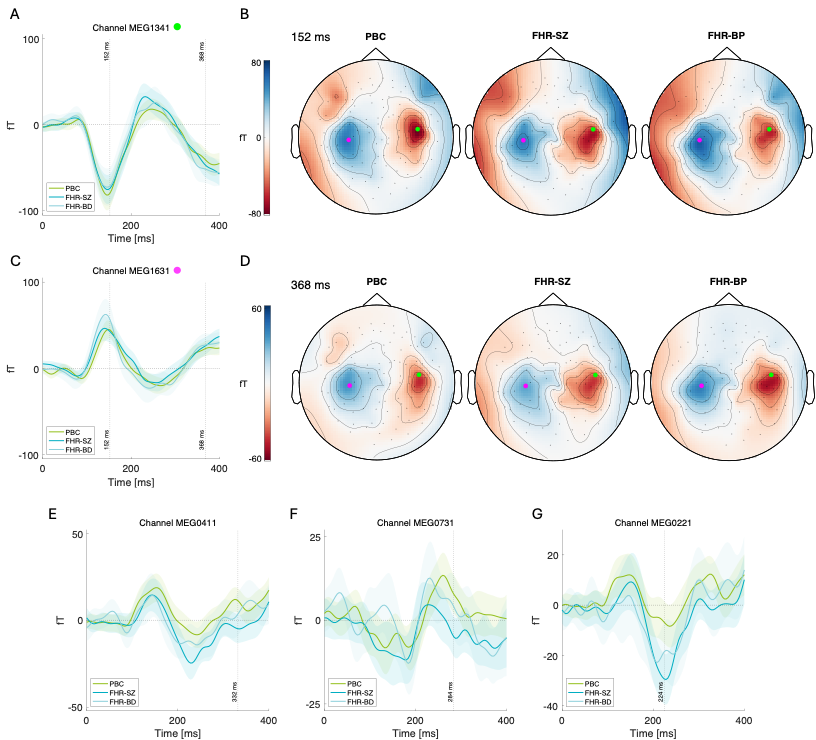

Supplementary Figure S3. Sensor-level event-related fields and scalp topographies by group. A) and C) display event-related fields for each group at sensors near maximal condition (oddball) effects in population-based controls (PBC). B) and D) display scalp topographies for each group at two time points (152 and 368 ms) for maximal condition (oddball) effects in PBCs, see Table S3 for peak coordinates and statistics. Green and magenta dots mark the location of the sensors displayed under panels A and C. Panels E-G display event-related fields for each group at sensors near the largest group differences (p < 0.001) between individuals at familial high-risk for schizophrenia (FHR-SZ) and PBCs, see Table S7 for peak coordinates and statistics. Vertical dotted lines mark peak effects (FHR-SZ vs. PBC) in time. FHR-BP = familial high-risk of bipolar disorder.

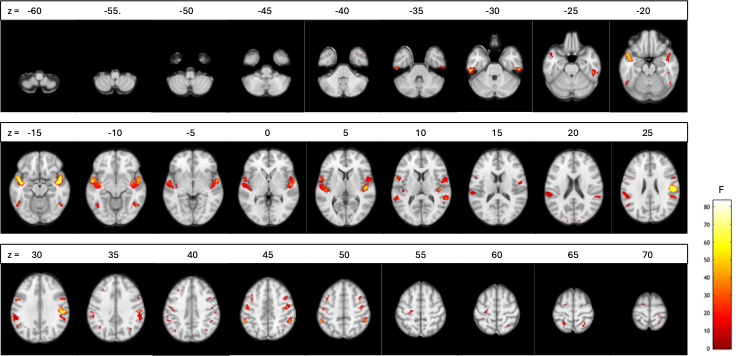

Supplementary Figure S4. Horizontal brain slices displaying differentially activated brain regions (deviants vs. standards, paired t-test, p < 0.05) in the total sample (n = 172).

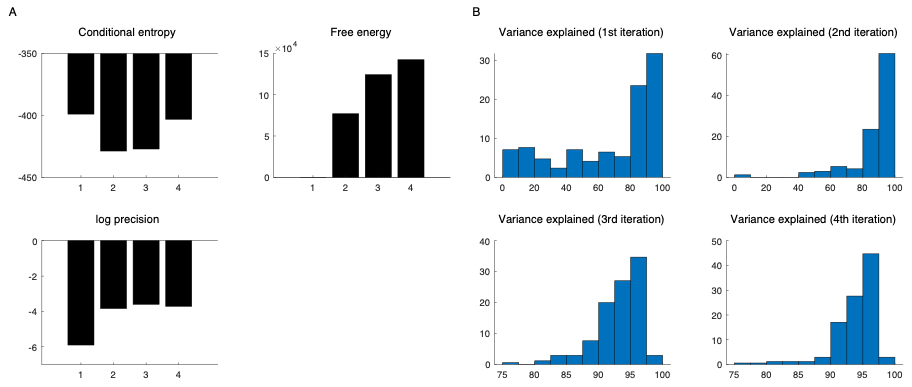

Supplementary Figure S5. Diagnostics for the iterative parametric empirical Bayesian (PEB) inversion. A) Conditional entropy, free energy, and log precision for each iteration (1-4). As visualised, the 2^nd^ iteration improved all three measures. The 3^rd^ iteration had minor effects on the log precision and conditional entropy, but increased the free energy. The 4^th^ iteration worsened the conditional entropy, increased the free energy further, and had little effect on the log precision. B) As a fourth measure, we plot the variance explained (R^2^) between the predicted and observed responses over the four iterations. Note the change in x-axis limits from the 2^nd^ to the 3^rd^ iteration. As expected, re-inverting the individual-level models using group-level posteriors as priors increased the variance explained, presumably due to the avoidance of local maxima. The increase was most pronounced from 1^st^ to 2^nd^ and from 2^nd^ to 3^rd^ iteration, with little improvement from 3^rd^ to 4^th^ iteration. The 3^rd^ iteration was chosen as the optimal, as it had the highest log precision (22) and was taken for further second-level analysis.

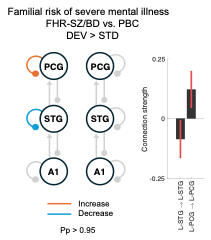

Supplementary Figure S6. Oddball-dependent effective connectivity in adolescents at familial high-risk of either schizophrenia or bipolar disorder (FHR-SZ/BP) versus population-based controls (PBC). Results from second-level parametric empirical Bayesian (PEB) analysis. DEV = deviants. STD = standards. PCG = precentral gyrus. STG = superior temporal gyrus. A1 = primary auditory cortex. Pp = posterior probability. L = left. R = right.
