## Supplementary material for "Magnetoencephalography reveals altered synaptic gain linked to functional impairment in adolescents at familial risk for schizophrenia and bipolar disorder": Table 1

**Table 1. Demographic and clinical characteristics of the participants.**

|  | PBC | FHR-SZ | FHR-BD | Total sample | p-value* | Pairwise comparisons, p-value** | | |
| --- | --- | --- | --- | --- | --- | --- | --- | --- |
|  |  |  |  |  |  | FHR-SZ  vs.  PBC | FHR-BP  vs.  PBC | FHR-SZ  vs.  FHR-BP |
| Participants, N | 75 | 59 | 38 | 172 |  |  |  |  |
| Females, N (%) | 40 (53%) | 25 (42%) | 21 (55%) | 86 (50%) | 0.35 |  |  |  |
| Age, mean (SD) | 15.9 (0.41) | 16.0 (0.35) | 16.0 (0.47) | 16.0 (0.40) | 0.64 |  |  |  |
| Structural MRI obtained, n (%) | 61 (81%) | 52 (88%) | 30 (79%) | 142 (83%) | 0.43 |  |  |  |
| PRS obtained, n (%) | 63 (84%) | 42 (71%) | 29 (76%) | 134 (78%) | 0.73 |  |  |  |
| C-GAS, median (IQR) | 80 (17) | 63 (30) | 72.5 (28) | 73 (27) | <0.001 | <0.001 | 0.043 | 0.069 |
| CBCL total score, median (IQR) | 9 (15) | 22 (32) | 15 (23) | 14 (24) | <0.001 | <0.001 | 0.006 | 0.341 |
| Primary caregiver PSP, median (IQR) | 83 (11) | 66.5 (25) | 62.5 (30) | 80 (26) | <0.001 | <0.001 | <0.001 | 0.498 |

CBCL = The Child Behavior Checklist. CGAS = Childrens Global Assessment Scale. FHR-BD = familial high risk of bipolar disorder. FHR-SZ = familial high risk of schizophrenia. IQR = interquartile range. PSP = Personal and Social Performance scale. SD = standard deviation.

Number of subjects with data: CGAS: n = 172. CBCL: n = 170. PSP = 172.

* P-values for significance testing using Pearson chi-squared for binary variables; analysis of variance (ANOVA) for normal continuous data; and Kruskal Wallis’s test for non-normal continuous variables (CBCL); all with high-risk status as the sole dependent variable.

** P-values for significance testing using t-tests (CGAS, PSP), and Wilcoxon ranksum tests for non-normal data (CBCL).
